# "We have our reasons": Exploring the acceptability of pre-exposure prophylaxis among gay, bisexual, and other men who have sex with men in Ghana

**DOI:** 10.1101/2023.12.05.23299515

**Authors:** Gloria Aidoo-Frimpong, Gamji Rabiu Abu-Ba’are, Amos Apreku, Richard Panix Amoh-Otu, Edem Zigah, Prince Amu-Adu, Samuel Amuah, Laura Nyblade, Kwasi Torpey, LaRon E. Nelson

## Abstract

Ghanaian men who have sex with men (MSM) face significant HIV disparities. Pre-exposure prophylaxis (PrEP) is a highly effective tool for HIV prevention. Previous studies on the perspectives of PrEP use among Ghanaian MSM identified high interest in PrEP among this population. However, the knowledge from the previous research, which was the best available evidence at the time, was primarily hypothetical because those data were collected before any real-world implementation of PrEP in Ghana. The purpose of the analysis is to identify and understand the factors currently influencing PrEP acceptance. We conducted a secondary analysis of focus group (n=8) data with Ghanaian MSM. Audio transcripts were subjected to descriptive thematic analysis. There was an almost universal awareness of PrEP, but inaccuracies about PrEP were common. PrEP acceptability was influenced by a mix of individual and intrapersonal factors. To bridge the gap between awareness, knowledge, and acceptability, HIV prevention programs should address access barriers and incorporate community-derived strategies.

## Introduction

Globally, HIV prevalence has decreased over the past decade, yet it remains disproportionately high among key populations, notably men who have sex with men (MSM). Key populations, including MSM, account for 70% of the global HIV burden, facing a risk 28 times greater than that of adult men who do not have sex with men (1). In sub-Saharan Africa alone, where 60% of the new 1.7 million HIV infections occurred in 2018 (2), Ghana presents a generalized HIV epidemic with prevalence rates 18.1% among MSM (3–5), compared to the 1.7% reported among the general population (6).

In response to this crisis, the global health community has embraced antiretroviral medications for pre-exposure prophylaxis (PrEP) as a preventive measure. The World Health Organization recommends PrEP as a highly effective biomedical prevention strategy for individuals at high risk of HIV exposure (7). The introduction of oral PrEP has been a significant advancement in the prevention of HIV (8), and more recently, the development of an injectable platform – which offers a sustained release formulation and was shown to be superior to oral PrEP in a randomized controlled trial, mainly due to its reduced reliance on daily adherence (9) – has broadened prevention strategies. Despite its promise, the uptake of PrEP is hindered by challenges at various levels, including provider, patient, and healthcare system (10, 11). The persistent barriers and lack of comprehensive knowledge among MSM significantly impede both the implementation and utilization of PrEP as an HIV prevention strategy.

In Ghana, the National AIDS/STI Control Program (NACP) under the auspices of the Ghana Health Service laid the groundwork for PrEP delivery by incorporating it into the Consolidated Guidelines for HIV Care in Ghana as early as August 2019 (12). This proactive step paved the way for the commencement of PrEP services to key populations a year later, in August 2020, with a particular emphasis on MSM (13). Research conducted in Ghana before the rollout of PrEP consistently indicated that a lack of awareness and knowledge among Ghanaian MSM was a significant barrier to PrEP’s acceptance(14, 15).

To maximize the effectiveness of PrEP in reducing HIV incidence among key populations in Ghana, particularly as access to this preventive measure expands, it is crucial to understand what factors influence both the awareness, knowledge and the acceptability of PrEP among Ghanaian MSM. This study’s primary goal was to explore the determinants of PrEP acceptability within a cohort of gay and bisexual men, as well as other men who engage in same-gender sexual relations in Ghana. Our research was driven by two principal questions: Firstly, what levels of knowledge, awareness, and attitudes do these men hold regarding PrEP? Secondly, do they view PrEP as an acceptable and efficient means of preventing HIV? Understanding the awareness, knowledge, attitudes, and acceptability of PrEP among MSM in Ghana is vital for the design of tailored educational and health promotion programs. This investigation aimed to reveal these factors to inform strategies that could bridge gaps in PrEP utilization. By identifying the current factors to PrEP uptake, this study could lead to the development of targeted interventions that enhance PrEP adherence and persistence, ultimately contributing to the reduction of new HIV infections among MSM in Ghana.

## Material and methods

This study was a qualitative description, using secondary data obtained from focus groups (FG) FGs are a type of group discussion that are used to elicit detailed descriptions of the shared experiences of those participating in the conversation(16). The social interaction that influences that discussion is a key advantage to understanding the general social perspective on a topic, which is not reliably achieved in an in-depth interview—which centers the individual’s perspective (17). These group discussions were conducted as part of formative research aimed at informing the adaptation of a multi-level intersectional stigma-reduction intervention. A comprehensive report on the recruitment methodology utilized in the parent study has been previously published (18). We provide a summary below.

### Study settings and sites

The parent study was conducted in Accra, the administrative capital, commercial hub, and largest city of Ghana, as well as in Kumasi, the second largest city in the country. These locations exhibit a rich blend of ethnicities, religions, and cultural diversity, including migrants from various regions within Ghana and neighboring countries. Further, these cities are located in the Greater Accra Region and Ashanti Region which were selected due to their high prevalence of HIV among MSM and their relatively larger populations of men with histories of same-gender sex, compared to other regions in Ghana.

### Study population and recruitment

Eligibility for participation in the FGs was limited to individuals who were 18 years of age or older, designated male at birth, currently self-identified as cisgender men, and reported engaging in sexual activity with another man (cisgender and transgender inclusive) at least once within the previous six months. Convenience sampling methods were utilized in partnership with two community partner organizations providing services to gender and sexual minorities to recruit study participants, who were recruited from September 1^st^, 2020 to October 30^th^, 2020.

### Focus Group Guides

The guides used for focus groups (FGs) contained open-ended questions covering ten main domains. The focus of the secondary analysis is on participants responses to the following questions used to stimulate discussion in the "Attitudes towards PrEP domain" Sample questions include (1) "Were you aware of PrEP before this study? (2) Describe what you know about PrEP, (3) If PrEP were available would you use it? (4) What will prevent or aid you in using PrEP for HIV prevention?

### Data Collection

The FG moderators involved in the study were individuals who self-identified as gay and bisexual men and were employees of LGBTQ+ serving partner organizations in their respective cities. These individuals were trained in data collection and the responsible conduct of research with human participants. A total of six FGs were conducted in English. Two FGs were conducted in Twi which is the predominant language spoken in Kumasi, which is the seat of the Twi-speaking Ashanti Kingdom. Providing participants with a non-English language option allowed for the narration of certain experiences that could only be effectively described in their local indigenous language. Participants were informed about the study, its voluntary nature, and the option to withdraw without consequences. Once a participant verbally indicated to the research assistant that all their questions had been satisfactorily answered, we documented that they had given informed consent by having the participant sign an informed consent form. FGs lasted for approximately 90-120 minutes. All FGs were digitally recorded and transcribed. The research team, including bilingual research assistants reviewed both English and Twi audio files to ensure consistency of translation and accuracy of the transcription.

### Data analysis

Seven research team members with diverse expertise and cultural backgrounds analyzed the data transcripts. The team began by individually analyzing six randomly selected transcripts from the FGs, then identifying and labeling relevant concepts. This was the first step in the coding process. The team then convened to construct a coding schema by defining and categorizing these individual codes. The team used the resultant schema to code the rest of the transcripts. Any discrepancies between coded text were discussed and addressed through discussion with the larger group of analysts and revision of the coding schema to include any new codes. We then combined all codes into code clusters. Clusters were then grouped together into broad categories of participants’ experiences and perspectives. The research team examined the resulting broad categories across all transcripts and reviewed them to confirm their comprehensiveness and representativeness. Finally, we identified themes by determining the main message(s) across the broad categories (19).

### Ethical Review

Approval for this study was obtained from the institutional review boards of Yale University, Noguchi Medical Research Institute (which serves as the IRB for the University of Ghana), the Ghana Health Service, and the University of Toronto. Before their participation, all study participants who agreed to participate signed an informed consent form. As a token of appreciation for their participation, study participants received a small gratuity of 100 Ghanaian Cedis (approximately $10 USD) for participating in focus group discussions.

## RESULTS

Two overarching themes were identified that characterized the men’s awareness, knowledge and acceptability of PrEP. Awareness was conceptualized as the conscious recognition of the existence of PrEP. Knowledge pertained to the accuracy of details about how it works and how it’s supposed to be used, including its contraindications (e.g., don’t take if you have a suspected HIV infection because of the risk of developing ARV resistance) and limits (e.g., doesn’t prevent STIs). Each of the two themes is built on four categories. The first theme, “almost universal awareness of PrEP but inaccuracies about PrEP were common” are underpinned by these categories: 1) confusion between pre-exposure prophylaxis and post-exposure prophylaxis, 2) event-driven perceptions of PrEP usage, 3) misaligned analogies: equating PrEP with ARVs, and 4) advocacy for a comprehensive educational approach. The second theme was PrEP acceptability was influenced by a mix of individual and intrapersonal factors. The corresponding categories were: 1) Hesitancy towards PrEP uptake due to perceived limited benefit, 2) Extending HIV, sex, and same-sex relations stigma to PrEP affects PrEP uptake, 3) Perceived lack of information, concerns about side effects, and cost act as barriers to PrEP acceptability. These identified themes highlight the importance of addressing gaps in awareness and knowledge of PrEP while acknowledging the complex and diverse factors that influence PrEP’s acceptability.

### Theme 1: There was an almost universal awareness of PrEP but inaccuracies about PrEP were common

#### Confusion between pre-exposure prophylaxis and post-exposure prophylaxis

All the participants in the FGs had heard of PrEP prior to the interview, with participants in two FGS indicating that they knew what PrEP was used for. Among those who indicated knowledge of PrEP, it was generally understood as a medication you take after exposure to HIV, thus describing post-exposure prophylaxis (PEP), and not PrEP, which is for pre-exposure. In the except below, a participant reveals a notable misconception surrounding PrEP (pre-exposure prophylaxis) and its confusion with post-exposure prophylaxis (PEP). The participant’s comment reflects a context where PrEP is associated with a specific use case – providing medication to staff who have been exposed to potential HIV transmission sources like blood or blood products.

> *"We use it for our staff, any staff who get infected, you get exposed to blood or blood products, PrEP or blood splash, we test you and give you PrEP then three months later we test you again, if you are fine, fine.* (Group 3, Accra).

This misunderstanding indicates that some individuals might perceive PrEP as a form of "emergency" or "post-exposure" treatment, rather than its intended use as a preventive measure before potential exposure. This perception could stem from inaccurate information dissemination. This highlights a crucial need for educational interventions to clarify the distinct purposes of PrEP and PEP among the target population. Additionally, it underlines the significance of accurate communication about PrEP’s pre-exposure nature to maximize its effectiveness in HIV prevention efforts.

#### Event-Driven Perceptions of PrEP Usage

In two focus groups in Kumasi, the men discussed their knowledge on PrEP, describing it as an event-driven prevention method. The perspectives shared underscored a mixture of awareness and misperception. As highlighted by a participant: “*What I know about PrEP is that you need to take it for about a month for prevention and after the month you take it when you are ready for sex*” (FG4, Kumasi). While correctly acknowledging the need to take PrEP for prevention, there was an apparent misunderstanding regarding the timing of its consumption. The idea of taking PrEP "for about a month" may stem from the initial period required for the medication to establish its protective effects. However, the belief that PrEP is only taken "when you are ready for sex" suggests a misconception about its continuous daily use, irrespective of sexual activity timing.

#### Misaligned Analogies: Equating PrEP with ARVs

Regarding the participants’ awareness of PrEP availability in Ghana, a consensus emerged that PrEP was indeed available within the country. However, an interesting perception was highlighted during the discussions – some participants drew parallels between PrEP and antiretroviral medications (ARVs). This analogy revealed a belief that, akin to ARVs, PrEP is tailored to individual needs. This sentiment was captured by a participant’s comment: "*As it is PrEP is now becoming common. PrEP is just not like paracetamol, it’s a variety of medications. The PrEP you will take will be different from the PrEP I may take.*" (Group 3 Accra) This perspective unveils a potential misinterpretation, as PrEP is generally standardized regardless of personal differences. This misconception underlines the significance of clarifying PrEP’s uniformity and its distinct nature from ARVs.

A group emphasized the necessity of simplifying the description of PrEP to enhance comprehension. This entailed equating PrEP to antiretroviral medications (ARVs) and offering a level of education comparable to that provided to individuals living with HIV. The sentiment was articulated by a participant who remarked, "*The thing is that, we need to admit that PrEP currently in Ghana is simply ARVs for all of us to understand. It’s the same medication that people living with HIV take. The only difference here is that, we use three different medicines for people who are HIV positive but if you are taking it for PrEP, we use two combinations, so instead of three medicines, if you want to take PrEP, you just take two. That is basically PrEP.*" (Group 4, Accra). Discussions highlighted the importance of providing clear, relatable information to dispel misconceptions and promote accurate comprehension among the population.

#### Advocacy for a Comprehensive Educational Approach

Participants emphasized the pressing need for intensified education to enhance PrEP awareness and understanding, acknowledging the prevalent lack of knowledge. The consensus was that delivering comprehensive information was pivotal, and should include all aspects such as dosage, potential side effects, long-term implications, and general information about PrEP including where to access it. Participants believed that increasing knowledge may lead to increased acceptability among MSM in Ghana. One participant shared: "*We talked about PrEP. I think the education on PrEP is not that strong, I think we should do more education on PrEP because it seems like people don’t understand and don’t know the reason why they will be taking drugs to prevent only HIV because I am a case manager, I test people who tend to be negative and I try to preach the gospel to them about PrEP but you can see that the community is not accepting it. I can see that the education is not that strong, so I think we have a lot to do about PrEP issue*" (Group 4, Accra). This perspective underscored the pivotal role of informative campaigns that dispelled misunderstandings, addressed concerns, and cultivated a deeper understanding of PrEP’s purpose and benefits. By bolstering education initiatives, the participants envisioned the potential to positively shape the community’s perception of PrEP and ultimately foster its wider acceptance and uptake.

### Theme 2: PrEP acceptability was influenced by a mix of individual and intrapersonal factors

#### Hesitancy towards PrEP uptake due to perceived limited benefit

In all eight FGs, participants were quite hesitant about PrEP uptake. The most recurring reason shared by participants was the limited benefit of PrEP in preventing HIV only and not other STIs. Participants felt that it was quite limiting to take a medication daily to prevent only one disease when usually the concern is more about pregnancy with their female partners and STIs such as gonorrhea and syphilis with their male partners. They described taking PrEP as an inconvenience, especially if they would need to take other actions to prevent other STIs. The statement from a participant: "*I cannot waste my time taking drugs every day to prevent me from HIV, only HIV, for me [to be] catching other STIs so it’s better for me to leave this PrEP and concentrate on my condom use*. (Group 4, Accra) presents a compelling perspective on the perceived drawbacks of using PrEP as a HIV prevention strategy. This discussion highlights several important insights. Firstly, it underscores the commitment required for PrEP use, as it involves daily medication adherence. For some individuals, this daily regimen may be perceived as inconvenient or unnecessary, particularly if they believe they are still at risk of contracting other sexually transmitted infections (STIs). This perception of inconvenience may deter them from utilizing PrEP consistently. Secondly, the discussions reflect concerns about the limited scope of PrEP in preventing only HIV. They expressed a preference for using condoms as a comprehensive prevention method against both HIV and STIs. This viewpoint aligns with the importance of emphasizing that PrEP is not a substitute for safer sex practices but rather a complement to them. It highlights the need for comprehensive sexual health education to clarify PrEP’s role in a broader prevention strategy. Finally, the discussions underline the fact that individuals have different risk perceptions and preferences when it comes to HIV prevention. Some may prioritize condom use due to its dual protection against HIV and STIs, while others may find PrEP more suitable for their lifestyle.

While the most frequent preference was not to use PrEP because it only protects against HIV, participants in one focus group in Kumasi differed. In this group, participants acknowledged the limitation of PrEP in protecting against other sexually transmitted infections (STIs). However, they introduced a unique perspective driven by their motivations for considering PrEP. The statement from Group 3 in Kumasi, "*We have our reasons though, we understand PrEP doesn’t prevent other STIs but at least you have HIV to not worry about,*" offers a nuanced perspective on the decision to adopt PrEP despite its limitations. This viewpoint sheds light on the trade-offs individuals may consider when making choices about their sexual health. Group 3’s acknowledgment that PrEP doesn’t provide protection against other STIs demonstrates a clear understanding of PrEP’s scope and limitations. However, their willingness to use PrEP primarily to prevent HIV suggests that they prioritize this particular concern above others. This choice reflects a calculated decision based on perceived risk and priorities. The phrase "*at least you have HIV to not worry about"* underscores the significance of HIV as a major health concern. For some individuals, the peace of mind that comes from knowing they are protected against HIV may outweigh the concern of contracting other STIs. It’s important to note that this perspective may vary among individuals, and their risk perceptions and priorities can influence their choices regarding prevention methods. The insights generated from this discussion highlights the complexity of sexual health decision-making and the importance of offering a range of prevention options that cater to individual preferences and risk perceptions.

There was strong preference for condom use overall as the preferred HIV prevention method. Condoms were characterized as a routine part of sexual encounters and remained the primary HIV prevention strategy they were most familiar with. Participants discussed condoms as convenient, and less burdensome compared with "*the stress of taking a pill every day*" (Group 1, Kumasi). Additionally, discussion revealed a firm decision against taking PrEP, underpinned by their current practices of using condoms and lubricants as preventive measures against both HIV and sexually transmitted infections (STIs). Their decision on emerged from their confidence in their existing practices, specifically the use of condoms and lubricants, as effective measures against both HIV and STIs. The collective sentiment, which was echoed by a participant, "*I will not take it. Since I am still using condoms and lubricant, I’m using them correctly and I am using them to prevent HIV and STI’s already, so I am not going to double protect myself.”* (Group 2, Accra) signifies their belief in their method’s sufficiency. This shared viewpoint reflected the group’s understanding of layered preventive strategies and their perception that adding PrEP would be redundant.

Within the discussions of two focus groups in Accra and one in Kumasi, a shared perspective emerged regarding the necessity of using condoms for protection, even in the presence of PrEP availability. Participants exhibited a degree of skepticism towards the standalone effectiveness of PrEP, suggesting that its protective capability might be insufficient when considered alone. This skepticism led participants to favor condoms as a more reliable option for ensuring their sexual health. As explained by this participant: "*I am just getting the information that even with the PrEP you still need to, there is a bigger possibility of you still using a condom, which means that PrEP doesn’t really do anything. Let me go for the condom and know I am safe"* (Group 1, Accra). This collective viewpoint underscores the participants’ cautious approach to preventive measures, revealing an inclination to adopt multiple strategies for added reassurance. The participants’ stance highlights the importance of addressing concerns about PrEP’s effectiveness and promoting comprehensive education about its intended role as part of a multi-faceted approach to HIV prevention.

#### Extending HIV, sex, and same-sex intercourse stigma to PrEP affects PrEP uptake

Stigma emerged as a notable barrier to PrEP acceptability, with a particular focus on the stigma associated with individuals who are living with HIV. The participants’ perspectives unveiled two main points. Firstly, a prevailing assumption arose that individuals taking PrEP were already living with HIV. Secondly, a perception existed that those living with HIV were subjected to stigma and discrimination. Despite recognizing the potential of PrEP to prevent HIV acquisition, participants expressed concerns that using PrEP might imply living with HIV. A participant succinctly shared: "*Trust me, if it [PrEP] is being advertised and made public for everyone to know in Ghana where we are, anyone who sees you holding the drug [PrEP] would say you are positive and won’t even come closer to you again*" (Group 2, Kumasi) This highlights the profound impact of HIV stigma on individuals’ perceptions of and willingness to use PrEP. It serves as a reminder of the ongoing challenges in combating stigma and the need for comprehensive strategies that not only provide access to prevention methods but also address the social and cultural factors that affect their adoption.

In all of the group discussions, there were concerns with being identified as living with HIV, an identity which is currently stigmatized. Participants were concerned about having a similar medication schedule with persons living with HIV, in which they both had to take a pill daily: " *I think there is no different the one who has HIV and the one who doesn’t have. This is because the one with HIV takes drugs everyday as well as the one who takes PrEP*" (Group 4, Accra). This raises an important perspective regarding the perceived similarity between individuals taking antiretroviral therapy (ART) for HIV treatment and those using PrEP for HIV prevention. This viewpoint suggests a fundamental misconception that exists in some communities, equating PrEP use with being a person living with HIV. It highlights the need for effective public health communication to distinguish between the two scenarios. While both individuals may take daily medications, their reasons for doing so differ significantly. ART is used to manage and control HIV in individuals who are already HIV-positive, while PrEP is a preventive measure for those at risk of HIV infection.

Moreover, the study identified a third reason for potential stigmatization—people assuming PrEP usage as indicative of engaging in "risky" sexual behaviors, primarily among men who have sex with men (MSM). As described by one participant: "*even in the hospital, to get a PrEP thing, it’s a ‘eiiii’ why do you need PrEP, why are you asking of PrEP, they have already perceived you to be going to do something so you need PrEP*" (Group 3, Accra). This perspective highlights a deep-seated bias and misunderstanding surrounding PrEP and the individuals who might benefit from it. The perception that merely inquiring about or seeking PrEP is met with judgment and suspicion indicates a prejudiced mindset prevalent in healthcare settings. This stereotype assumes that anyone interested in PrEP must already be engaging in behaviors that put them at risk for HIV, which can lead to discrimination and stigmatization. This type of stigmatization poses a significant barrier to HIV prevention efforts, particularly among vulnerable populations like MSM. It discourages individuals from seeking the necessary healthcare services, including PrEP, due to fear of being unfairly judged or stigmatized by healthcare providers and the broader community.

Discussions among the groups emphasized a complex web of stigma and barriers associated with seeking PrEP in Ghana. One significant concern raised by focus groups is the fear that seeking PrEP within healthcare settings may inadvertently lead to self-identification as an LGBTQ+ individual, which can further contribute to stigmatization. As described by a participant: “*In government hospital, it will take a lot of push and pull before we will give it to you, even that, they will ask, are you MSM?*” (Group 3, Accra). The highlights the challenging dynamics within healthcare settings. The requirement to visit a hospital to access PrEP can be viewed as a source of stigma itself, as it may expose individuals to judgment and scrutiny. The mention of healthcare providers asking, "*are you MSM?"* underscores the added layer of stigma faced by LGBTQ+ individuals. This line of questioning can make individuals feel uncomfortable, potentially leading them to withhold information about their sexual orientation or behaviors, which is detrimental to their overall healthcare. Furthermore, the fear of being publicly identified as a PrEP user and having one’s sexual identity and behaviors recorded can be a significant deterrent to accessing care, especially in environments where same-gender sexual relationships are criminalized. This concern is rooted in the very real fear of legal repercussions and societal discrimination faced by LGBTQ+ individuals in such context.

A prevailing sentiment across the groups was the apprehension that introducing PrEP into the MSM community might inadvertently encourage a surge in high-risk sexual behavior and the development of multiple sexual partnerships. These concerns, voiced by all but one focus group (Group 1, Kumasi) revolved around the potential negative consequences such behavior might entail within the MSM community. Participants expressed reservations that the availability of PrEP, while aiming to prevent HIV among individuals engaging in risky behaviors, could inadvertently promote such behaviors by providing a sense of protection. A participant’s perspective, "*Since PrEP prevents HIV in people who live risky behaviors, thus encouraging people to live that risky lifestyle and this will prevent me from using it*" (Group 3, Kumasi) succinctly captures this sentiment. The apprehension expressed by the men underscores the intricate relationship between preventive measures like PrEP and individuals’ behavioral patterns. It highlights the importance of education and communication strategies that clarify PrEP’s role within a comprehensive sexual health strategy and address concerns about its potential impact on behavior.

#### Perceived lack of information, concerns about side effects, and cost act as barriers to PrEP acceptability

Amid the various themes discussed, a common thread of confusion and insufficient information about PrEP dosing, processes, and side effects emerged across all focus groups. While not as widespread as other concerns, this lack of clarity was evident. Participants consistently expressed a sense of inadequate awareness surrounding PrEP, with misconceptions particularly noticeable in relation to dosing schedules and potential ramifications of missed dosages. A participant’s question, "*Does that mean that I may contract the virus when I fail to take PrEP even for a day?*" (Group 4, Kumasi), exemplifies this uncertainty. This prevailing sentiment highlights the critical need for comprehensive educational campaigns that provide accurate and transparent information about PrEP, addressing its dosing regimens and clarifying misconceptions regarding missed doses. The participants’ shared confusion underlines the importance of cultivating an informed population that can make sound decisions about PrEP usage, maximizing its effectiveness as an HIV prevention strategy.

The participants consistently highlighted concerns regarding potential side effects, particularly those related to kidney health, as well as perceived complications associated with PrEP usage. The participants’ apprehension was rooted in the fear that the medication could exacerbate pre-existing health conditions, causing complications. A participant’s remark, "*With the PrEP I think, if you have some kind of underlying illnesses and all of that, it would give you complications*" (Group 1, Kumasi), encapsulates this viewpoint. The participants’ anxieties were further reinforced by the knowledge that PrEP might impact kidney function, as indicated by another participant’s statement, "*Knowing right now that it affects the kidney, no*" (Group 4, Accra). This shared apprehension unveils a significant information gap that needs to be addressed during education campaigns and interventions. The participants’ collective perspective underscores the importance of providing transparent and accurate information about PrEP’s potential side effects, its impact on existing health conditions, and the specific medical considerations that individuals should take into account before opting for PrEP.

The issue of cost was a concern raised by participants in one FGD (Kumasi), who were worried that PrEP might be prohibitively expensive. Participants voiced worries that the expense associated with PrEP could render it financially inaccessible for many individuals. The perceived high cost of PrEP raised fears that it might pose a substantial barrier to access, potentially preventing a considerable portion of the population from benefiting from the medication. A participant succinctly captured this sentiment with the statement, "*The price may scare me from using it*" (Group 1, Kumasi). The participants’ collective viewpoint underscores the importance of addressing cost-related apprehensions during awareness campaigns and intervention strategies. It emphasizes the necessity of providing clear information about the affordability of PrEP, including potential subsidies or cost-reduction mechanisms, to ensure that individuals are well-informed and empowered to make decisions based on accurate information rather than financial constraints.

#### Varied reasons for participant acceptability of PrEP

Amid these findings, select focus groups (three) highlighted potential determinants that could influence PrEP acceptability. Participants’ motivations for considering PrEP usage encompassed various factors, including HIV prevention, addressing Hepatitis B concerns, maintaining intimacy in serodiscordant relationships, and navigating issues of trust within partnerships. Participants engaged in discussions regarding their readiness to embrace PrEP for HIV prevention, centering their considerations on personal safety and a strong desire to remain HIV-free. The conversations also underscored the added advantage of PrEP in treating Hepatitis B, which contributed to participants’ inclination to opt for PrEP. This perspective is succinctly expressed by a participant’s comment, "*the PrEP medication also treats Hepatitis B alongside, So I will use it*" (Group 1, Accra). Additionally, participants echoed their willingness to adopt PrEP, emphasizing its role in averting HIV transmission: "*Yes, I will use it because it will help me prevent HIV*" (Group 4, Kumasi). This sentiment was reinforced by participants who viewed PrEP as an additional safety measure: "*I think I would like to use it just to be safe*" (Group 2, Kumasi). These shared viewpoints illuminate the multifaceted considerations that influence PrEP acceptability. They underline the significance of a holistic approach to PrEP awareness and education that addresses various motivating factors, including personal health safety and the potential for treating co-occurring conditions like Hepatitis B, ultimately contributing to informed and comprehensive HIV prevention decisions.

Several participants voiced a perspective that PrEP holds substantial value for HIV serodiscordant couples, as it enables a level of sexual intimacy that might otherwise be hindered by concerns about HIV transmission. This concern is particularly significant when the uninfected partner is uncertain about their partner’s viral suppression or treatment adherence. These participants emphasized their anxieties about their partner’s fidelity and commitment to monogamy. In response, they expressed a willingness to utilize PrEP as a means of self-protection against potential transmission risks. A participant explained: *"I would like to use it because being with my “beef” and not knowing whether he is faithful, I would love to take it just to prevent myself from anything*" (Group 2, Accra). This viewpoint reflected a complex interplay between sexual health, trust within relationships, and PrEP acceptability. The participants’ shared perspective highlights the role of PrEP in facilitating open communication about trust, fidelity, and safety within partnerships. It also underscores the significance of providing tailored support and information to couples navigating serodiscordant relationships, ultimately promoting the holistic well-being of both partners.

## DISCUSSION

PrEP is a proven effective method for preventing HIV acquisition, particularly for individuals with high odds of being exposed to the virus(20). Although PrEP was introduced in Ghana and other West African countries in recent years, there exists a limited understanding of its acceptability among Ghanaian MSM. This article, therefore, provides qualitative insights into awareness, knowledge, and perspectives of the acceptability of PrEP uptake among MSM in Ghana. The findings will contribute to shaping interventions to increase their PrEP uptake and ultimately reduce HIV incidence in the country. In this study, we observed that 1) There was an almost universal awareness of PrEP, but inaccuracies about PrEP were common, and 2) PrEP acceptability was influenced by a mix of individual and intrapersonal factors.

Our previous research in Ghana found a low awareness of PrEP among MSM(15). Our current results after the introduction of PrEP in Ghana indicate an almost universal awareness of PrEP among MSM. However, like other studies elsewhere, we observed a misunderstanding of the basics of PrEP, including its purpose, dosage, and effects (21–24). For instance, in explaining their reasons for taking or not taking PrEP, many participants equated PrEP to PEP, indicating that PrEP is taken after exposure (which is PEP in true definition). They also described that there is no difference between PrEP and ARVs as they are both daily pills; however, while oral PrEP is based on the daily use of an ARV combination, it is not a treatment for HIV. Additionally, their explanations suggest that they heightened fears of harm from taking PrEP medication, for example side effects such as impaired kidney functions and complications associated with pre-existing conditions. Whereas no other studies exist on PrEP among MSM after it was introduced in Ghana, our findings corresponds with a study among sex workers in Ghana, that show that low knowledge of PrEP affected interest in its uptake (14).

Similar to Ahouada et al.’s (25) finding among MSM in Benin, MSM in this study held mixed individual and intrapersonal perspectives that inform the acceptability or unacceptability of PrEP. For those who accepted to take PrEP, they pointed to critical issues regarding known HIV risk behaviors such as multiple sex partnerships, low condom use, and general risk behaviors among MSM and sexual partners. As such, MSM find PrEP a vital prevention option, as it will prevent them from contracting HIV even if their sexual partners are living with HIV. Those who indicated a lack of acceptability of PrEP cited factors such as the daily uptake of PrEP as inconvenient and not different from someone living with HIV and that PrEP does not prevent any other condition besides HIV. Hence, they do not think it’s worth the inconvenience.

Participants demonstrated a preference for traditional prevention methods like condom use, which they perceive as more accessible and straightforward compared to PrEP. The convenience of condoms, their effectiveness against a broad spectrum of STIs, and the immediacy of their protection are factors that underscore their continued favorability within the MSM community. This preference is further rooted in the practical challenges associated with PrEP, such as the daily commitment to medication adherence and confusion overdosing schedules, which could potentially diminish its preventive efficacy if doses are missed. These concerns reflect a broader need for clear, practical guidance on PrEP use to ensure that it becomes a more user-friendly option for those at high risk of HIV exposure.

There is an opportunity explore the use of injectable PrEP in Ghana as it has the potential to address the desire for a more convenient option that did not require daily dosing among MSM. It also possible that despite the concerns of side effects, MSM will still take PrEP if its injectable because of convenience. A qualitative study among men in Los Angeles in the US reported that despites many participants experiencing side effects, they found injectable PrEP more convenient to use as they do not have to worry about taking pills daily and also the advantage of not worrying about adherence to PrEP (26).

A noteworthy issue is the extension of HIV stigma, medication stigma, and sexual stigma to PrEP and the potential for such stigmas to affect PrEP uptake (27). The participants in this study indicated not taking PrEP because it will be equated to HIV treatment if others see it. Hence, others will avoid them because they assume they are living with HIV, thus expressing a fear (anticipation) of HIV stigma if using PrEP, which affects willingness to take PrEP. Some participants do not want to be associated with PrEP because it will be assumed that they are engaging in same-gender sex, especially if they must go to a healthcare setting for PrEP. Similar to reasons provided in our previous studies for not testing for HIV in healthcare facilities (28), participants expressed fear of disclosing their sexual behavior at healthcare facilities during PrEP enrolment. The fear of disclosure is expected as many Ghanaian MSM have expressed strong reservations about confidentiality in facilities in several studies (28–32). This echoes previous finding elsewhere that stigma and discrimination associated with HIV and sexuality plays a significant barrier to PrEP uptake among MSM (33, 34). For instance, a study conducted in West Africa found that societal attitudes towards homosexuality significantly hindered PrEP uptake among MSM (35). Similarly, a study in Thailand found that cultural norms and taboos surrounding same-sex behavior were significant barriers to PrEP uptake among MSM(36). Addressing these complex issues is essential for improving PrEP awareness and uptake among MSM in Ghana.

The participants highlighted the importance of upscaling comprehensive PrEP education among MSM in Ghana that will tackle areas such as dosage, side effects, and general information about PrEP. They recognized the lack of understanding of PrEP among themselves and other Ghanaian men who share similar sexual histories, despite its potential to reduce HIV acquisition. Thus, supporting previous calls for the scale up of interventions to address PrEP uptake among men who may be exposed to HIV through sexual contact with other men(37–40). Although research on interventions to promote PrEP use among MSM is limited, recent studies have explored the use of social networks to improve PrEP initiation and other forms of HIV prevention in this population. These interventions have aimed to leverage the power of social networks to increase awareness, promote education and reduce barriers to accessing PrEP and other forms of HIV prevention (39, 40). We encourage researchers and HIV prevention program implementers to explore these approaches, considering that it has been successful in previous studies in reaching MSM for intersectional stigma, HIV prevention, and testing interventions in Ghana (32, 41–43) and has been successful in contributing to an increase in PrEP uptake elsewhere (44, 45).

### Strengths and Limitations

A major strength of this study is its pioneering role in examining the knowledge, awareness, and acceptability of PrEP among MSM in Ghana following its introduction. This timely research fills a critical gap in the literature by providing fresh insights into the perceptions and experiences of this key population at a significant juncture in the local HIV prevention landscape. The qualitative design allows for an in-depth exploration of individual and collective attitudes toward PrEP, capturing the complexities and subtleties of these perceptions. Engagement with MSM through advocacy organizations offers informed perspectives that may indicate a higher baseline understanding of PrEP-related issues. The study’s limitations, however, must be acknowledged. It focuses solely on oral PrEP, overlooking the nascent but potentially transformative role of injectable PrEP, which could offer alternative adherence models and convenience. The recruitment strategy, centered on connections with advocacy organizations, may limit the representativeness of the sample, potentially excluding the voices of those with less support or awareness about PrEP.

With a modest focus group size and a lack of comprehensive sociodemographic data, the ability to extrapolate these findings to the wider MSM population in Ghana is limited. Furthermore, the absence of a quantitative measure constrains the ability to assess the prevalence of the observed attitudes and knowledge gaps, which would be beneficial for public health planning and intervention design. Given the recency of PrEP’s introduction in Ghana, the study does not capture longitudinal data, which would provide insights into the evolution of PrEP knowledge and attitudes over time, as well as the impact of ongoing educational efforts.

## Conclusions

PrEP stands as a pivotal intervention for the prevention of HIV, with clear efficacy for those at substantial risk of contracting the virus. Its introduction in Ghana and other regions of West Africa marks a crucial step in broadening HIV preventive measures. However, our understanding of its acceptability among Ghanaian men who have sex with men (MSM) remains nascent. This study contributes qualitative insights into the awareness, knowledge, and acceptability of PrEP among this group. The increased understanding gained here is instrumental in shaping targeted interventions to enhance PrEP uptake, which could play a significant role in reducing HIV incidence in Ghana. In light of these findings and limitations, future research should include an examination of the acceptability and feasibility of injectable PrEP among MSM in Ghana. As the healthcare landscape evolves to potentially include injectable options, understanding preferences and barriers to this modality will be crucial. This knowledge could inform the development of tailored interventions that accommodate the specific needs and preferences of this high-risk population, ultimately contributing to a more effective HIV prevention strategy in Ghana and similar contexts.

## Data Availability

The datasets used and/or analyzed during the current study are not publicly available due to our ethical and legal requirements related to protecting participant privacy and current ethical institutional approvals but are available from the corresponding author on reasonable request pending ethical approval.

## Acknowledgements

This study would not have been possible without the generosity of the study participants and their willingness to share their knowledge and experiences with us, and the expertise of our partner organizations and their staff: Priorities on Rights and Sexual Health (PORSH) in Accra, Youth Alliance for Health and Human Rights (YAHR), in Kumasi, and Educational Assessment Research Centre (EARC) in Accra. This study is sponsored by the National Institute of Nursing Research R01 NR019009, which provides direct financial support for the research. The study is also made possible through core services and support from the Yale Center for Interdisciplinary Research on AIDS (P30 MH06224), which provides ongoing consultation on research design and methods, including on methods to facilitate continuity of HIV/AIDS research operations during the COVID-19 pandemic. The Yale AIDS Prevention Training program (T32 MH020031) provides postdoctoral trainees to assist with research project coordination.

## References

1. UNAIDS. Global HIV & AIDS statistics — Fact sheet 2023 [Available from: https://www.unaids.org/en/resources/fact-sheet.

2. UNAIDS. World AIDS Day Report Geneva2018 [Available from: https://www.unaids.org/en/knowledge_is_power/share-graphics.

3. Aberle-Grasse J, McFarland W, El-Adas A, Quaye S, Atuahene K, Adanu R, et al., editors. HIV prevalence and correlates of infection among MSM: 4 areas in Ghana, the Ghana Men’s Health Study 2010-2011. 20th Conference on Retroviral and Opportunistic Infections (CROI 2013), Atlanta, GA; 2013.

4. Ghana AIDS Commission. GHANA MEN’S STUDY : Mapping and population size estimation (MPSE) and integrated bio-behavioral surveillance survey (IBBSS) amongst men who have sex with men (MSM) in Ghana (Round II). Accra. Ghana. 2017.

5. Phaswana-Mafuya N, Simbayi L, Wabiri N, Cloete A, Commission GA. The Ghana men’s study II: mapping and population size estimation (MPSE) and integrated bio-behavioral surveillance survey (IBBSS) amongst men who have sex with men (MSM) in Ghana. 2020.

6. Ghana AIDS Commission. GHANA’S HIV FACT SHEET 2021 2021 [Available from: https://www.ghanaids.gov.gh/.

7. World Health Organization. Pre-exposure prophylaxis (PrEP) 2021 [Available from: https://www.who.int/teams/global-hiv-hepatitis-and-stis-programmes/hiv/prevention/pre-exposure-prophylaxis#:~:text=As%20of%20September%202015%2C%20WHO,drug%20is%20used%20as%20directed.

8. Grant RM, Anderson PL, McMahan V, Liu A, Amico KR, Mehrotra M, et al. An observational study of preexposure prophylaxis uptake, sexual practices, and HIV incidence among men and transgender women who have sex with men. The Lancet Infectious diseases. 2014;14(9):820.

9. Landovitz RJ, Donnell D, Clement ME, Hanscom B, Cottle L, Coelho L, et al. Cabotegravir for HIV prevention in cisgender men and transgender women. New England Journal of Medicine. 2021;385(7):595–608.

10. Cowan FM, Delany-Moretlwe S, Sanders EJ, Mugo NR, Guedou FA, Alary M, et al. PrEP implementation research in Africa: what is new? Journal of the International AIDS Society. 2016;19:21101.

11. Mayer KH, Agwu A, Malebranche D. Barriers to the Wider Use of Pre-exposure Prophylaxis in the United States: A Narrative Review. Advances in Therapy. 2020;37(5):1778–811.

12. Ghan Health Service, National AIDS Control Programme. CONSOLIDATED GUIDELINES FOR HIV CARE IN GHANA: Test, Treat & Track. 2022.

13. Success Story: EpiC and Partners Introduce Pre-Exposure Prophylaxis in Ghana [press release]. 2021.

14. Guure C, Afagbedzi S, Torpey K. Willingness to take and ever use of pre-exposure prophylaxis among female sex workers in Ghana. Medicine. 2022;101(5).

15. Ogunbajo A, Leblanc NM, Kushwaha S, Boakye F, Hanson S, Smith MDR, et al. Knowledge and Acceptability of HIV pre-exposure prophylaxis (PrEP) among men who have sex with men (MSM) in Ghana. AIDS Care. 2020;32(3):330–6.

16. Morrison-Beedy D, Côté-Arsenault D, Feinstein NF. Maximizing results with focus groups: moderator and analysis issues. Appl Nurs Res. 2001;14(1):48–53.

17. Côté-Arsenault D, Morrison-Beedy D. Maintaining your focus in focus groups: avoiding common mistakes. Res Nurs Health. 2005;28(2):172–9.

18. Nelson LE, Nyblade L, Torpey K, Logie CH, Qian H-Z, Manu A, et al. Multi-level intersectional stigma reduction intervention to increase HIV testing among men who have sex with men in Ghana: Protocol for a cluster randomized controlled trial. PloS one. 2021;16(11):e0259324-e.

19. Morse JM. Confusing categories and themes. Qual Health Res. 2008;18(6):727–8.

20. Centers for Disease Control and Prevention. PrEP for HIV Prevention in the U.S. 2023

21. Aidoo-Frimpong G, Orom H, Agbemenu K, Collins RL, Morse GD, Nelson LE. Exploring Awareness, Perceptions, and Willingness to Use HIV Pre-Exposure Prophylaxis: A Qualitative Study of Ghanaian Immigrants in the United States. AIDS Patient Care STDS. 2022;36(1):8–16.

22. Yu S, Cross W, Lam LLY, Zhang K, Banik B, Li X, et al. Willingness, preferred ways and potential barriers to use pre-exposure prophylaxis for HIV prevention among men who have sex with men in China. BMJ open. 2021;11(10):e053634.

23. Iniesta C, Álvarez-del Arco D, García-Sousa LM, Alejos B, Díaz A, Sanz N, et al. Awareness, knowledge, use, willingness to use and need of Pre-Exposure Prophylaxis (PrEP) during World Gay Pride 2017. PLoS One. 2018;13(10):e0204738.

24. Kahle EM, Sullivan S, Stephenson R. Functional knowledge of pre-exposure prophylaxis for HIV prevention among participants in a web-based survey of sexually active gay, bisexual, and other men who have sex with men: Cross-sectional study. JMIR public health and surveillance. 2018;4(1):e8089.

25. Ahouada C, Diabaté S, Mondor M, Hessou S, Guédou FA, Béhanzin L, et al. Acceptability of pre-exposure prophylaxis for HIV prevention: facilitators, barriers and impact on sexual risk behaviors among men who have sex with men in Benin. BMC Public Health. 2020;20(1):1267.

26. Kerrigan D, Mantsios A, Grant R, Markowitz M, Defechereux P, La Mar M, et al. Expanding the menu of HIV prevention options: a qualitative study of experiences with long-acting injectable cabotegravir as PrEP in the context of a phase II trial in the United States. AIDS and Behavior. 2018;22:3540–9.

27. Dubov A, Galbo P, Jr., Altice FL, Fraenkel L. Stigma and Shame Experiences by MSM Who Take PrEP for HIV Prevention: A Qualitative Study. Am J Mens Health. 2018;12(6):1843–54.

28. Zigah EY, Abu-Ba’are GR, Shamrock OW, Dakpui HD, Apreku A, Boyd DT, et al. “For my safety and wellbeing, I always travel to seek health care in a distant facility”—the role of place and stigma in HIV testing decisions among GBMSM–BSGH 002. Health Place. 2023;83:103076.

29. Kushwaha S, Lalani Y, Maina G, Ogunbajo A, Wilton L, Agyarko-Poku T, et al. “But the moment they find out that you are MSM…”: a qualitative investigation of HIV prevention experiences among men who have sex with men (MSM) in Ghana’s health care system. BMC Public Health. 2017;17(1):770.

30. Ogunbajo A, Kershaw T, Kushwaha S, Boakye F, Wallace-Atiapah N-D, Nelson LE. Barriers, motivators, and facilitators to engagement in HIV care among HIV-infected Ghanaian men who have sex with men (MSM). AIDS and Behavior. 2018;22(3):829–39.

31. Saalim K, Amu-Adu P, Amoh-Otu RP, Akrong R, Abu-Ba’are GR, Stockton MA, et al. Multi-level manifestations of sexual stigma among men with same-gender sexual experience in Ghana. BMC Public Health. 2023;23(1):166.

32. Nyblade L, Stockton MA, Saalim K, Rabiu Abu-Ba’are G, Clay S, Chonta M, et al. Using a mixed-methods approach to adapt an HIV stigma reduction intervention to address intersectional stigma faced by men who have sex with men in Ghana. Journal of the International AIDS Society. 2022;25:e25908.

33. Chakrapani V, Newman PA, Shunmugam M, Mengle S, Varghese J, Nelson R, et al. Acceptability of HIV pre-exposure prophylaxis (PrEP) and implementation challenges among men who have sex with men in India: a qualitative investigation. AIDS patient care and STDs. 2015;29(10):569–77.

34. Jackson GY, Darlington CK, Van Tieu H, Brawner BM, Flores DD, Bannon JA, et al. Women’s views on communication with health care providers about pre-exposure prophylaxis (PrEP) for HIV prevention. Cult Health Sex. 2021:1–16.

35. Eubanks A, Coulibaly B, Dembélé Keita B, Anoma C, Dah TTE, Mensah E, et al. Socio-behavioral correlates of pre-exposure prophylaxis use and correct adherence in men who have sex with men in West Africa. BMC Public Health. 2022;22(1):1832.

36. Yang D, Chariyalertsak C, Wongthanee A, Kawichai S, Yotruean K, Saokhieo P, et al. Acceptability of pre-exposure prophylaxis among men who have sex with men and transgender women in Northern Thailand. PloS one. 2013;8(10):e76650.

37. Hotton AL, Keene L, Corbin DE, Schneider J, Voisin DR. The relationship between Black and gay community involvement and HIV-related risk behaviors among Black men who have sex with men. Journal of gay & lesbian social services. 2018;30(1):64–81.

38. Patel RR, Chan PA, Harrison LC, Mayer KH, Nunn A, Mena LA, et al. Missed Opportunities to Prescribe HIV Pre-Exposure Prophylaxis by Primary Care Providers in Saint Louis, Missouri. LGBT Health. 2018;5(4):25–256.

39. Quinn KG, Christenson E, Spector A, Amirkhanian Y, Kelly JA. The influence of peers on PrEP perceptions and use among young black gay, bisexual, and other men who have sex with men: a qualitative examination. Archives of sexual behavior. 2020;49:2129–43.

40. Quinn KG, Zarwell M, John SA, Christenson E, Walsh JL. Perceptions of PrEP use within primary relationships among young black gay, bisexual, and other men who have sex with men. Archives of Sexual Behavior. 2020;49:2117–28.

41. Nelson LE, Ogunbajo A, Abu-Ba’are GR, Conserve DF, Wilton L, Ndenkeh JJ, et al. Using the implementation research logic model as a lens to view experiences of implementing HIV prevention and care interventions with adolescent sexual minority men—A global perspective. AIDS and Behavior. 2023;27(Suppl 1):128–43.

42. Abubakari GMR, Owusu-Dampare F, Ogunbajo A, Gyasi J, Adu M, Appiah P, et al. HIV Education, Empathy, and Empowerment (HIVE 3): A Peer Support Intervention for Reducing Intersectional Stigma as a Barrier to HIV Testing among Men Who Have Sex with Men in Ghana. International journal of environmental research and public health. 2021;18(24):13103.

43. Abubakari GMR, Turner D, Nelson LE, Odhiambo AJ, Boakye F, Manu A, et al. An application of the ADAPT-ITT model to an evidence-based behavioral HIV prevention intervention for men who have sex with men in Ghana. International Health Trends and Perspectives. 2021;1(1):1–16.

44. Walters SM, Platt J, Anakaraonye A, Golub SA, Cunningham CO, Norton BL, et al. Considerations for the design of pre-exposure prophylaxis (PrEP) interventions for women: lessons learned from the implementation of a novel PrEP intervention. AIDS and Behavior. 2021;25(12):3987–99.

45. Newman PA, Akkakanjanasupar P, Tepjan S, Boborakhimov S, van Wijngaarden JWdL, Chonwanarat N. Peer education interventions for HIV prevention and sexual health with young people in Mekong Region countries: a scoping review and conceptual framework. Sexual and Reproductive Health Matters. 2022;30(1):2129374.

